# Effectiveness of aerobic physical exercise on depression symptoms in adults: A protocol for a systematic review and meta-analysis of randomized clinical trials

**DOI:** 10.1101/2024.11.19.24317605

**Authors:** Larissa Nayara de Souza, Silvana Medeiros de Araújo, Eva da Silva Paiva, Alícia Eliege da Silva, Joel Florêncio da Costa Neto, Juvêncio César Lima Assis, Isis Kelly dos Santos, Themis Cristina Mesquita Soares, Edson Fonseca Pinto, Roque Ribeiro da Silva Júnior, Maria Irany Knackfuss

## Abstract

**Objective:** Depression is a chronic condition that affects millions of people and requires effective interventions. Studies suggest that aerobic exercise can improve mental health and reduce depression symptoms, despite variations in exercise type and intensity. The aim of this study will be to analyze the effect of aerobic exercise on reducing depression in adults.

**Methods:** This is a systematic review protocol, which will follow the PRISMA-P 2020 guidelines and has been registered in PROSPERO (no. CRD42024592700). The study search will be conducted in five databases: PubMed, Embase, Cochrane, BVS, and SPORTDiscus, using MeSH-based descriptors. Studies will be selected independently by two researchers using the Rayyan ‘QCRI’ software. Data extraction will be conducted with specific forms, and the methodological quality of the studies will be assessed using the Cochrane RoB 2.0 method. The certainty of evidence will be evaluated through the Grading of Recommendations Assessment, Development, and Evaluation (GRADE).

**Conclusions:** The systematic review is expected to identify the effective dose-response for reducing depression levels and provide understanding of the mechanisms through which aerobic exercise influences depression.

## Introduction

According to the World Health Organization (WHO), more than 300 million people of all ages suffer from depression, also known as major depressive disorder (MDD) [1]. This situation highlights the urgency of finding effective strategies to address this serious public health issue.

In this context, physical exercise emerges as a promising intervention for improving mental health. Scientific evidence indicates that individuals with psychiatric disorders often exhibit compromised aerobic capacity and a high propensity for metabolic syndrome. Regular physical exercise not only helps mitigate these risk factors, but also promotes mental health and overall well-being [2].

Despite the documented benefits of aerobic exercise, the literature on this subject is vast and varied, with studies differing in exercise type, intensity, frequency, volume, and duration. Although there are indications that this type of exercise can effectively reduce depression levels, the underlying mechanisms of these effects are still not fully understood [3].

Therefore, this study is justified by the need to deepen understanding of how different intensities and types of aerobic exercise can influence reducing depression levels. The article aims to evaluate the effect of aerobic exercise on reducing depression levels in adults, based on results obtained from clinical trials investigating this condition by conducting a systematic review of randomized clinical trials.

## Methods

The study is a protocol for a systematic review, which will follow the guidelines developed by Page et al. [4] known as the Preferred Reporting Items for Systematic Reviews and Meta-Analyses (PRISMA-P) for systematic reviews and has been registered in the International Prospective Register of Systematic Reviews - PROSPERO, number: CRD42024592700.

### Research question

Thus, the PICOS clinical question was formulated as follows: Population: adults; Intervention: aerobic exercise; Comparator: other types of exercises/placebo/control; Outcome: improvement in depressive symptoms; Study types: randomized clinical trials (RCTs).

### Search strategy

The search will be conducted in the following databases: PubMed, Embase, Cochrane Library, Virtual Health Library (VHL), and SPORTDiscus. Additionally, Clinical Trials, Google Scholar, and the references of included studies will be scanned to identify potential studies.

The search strategy will be highly sensitive and systematic, using the official descriptors from the Medical Subject Headings (MeSH) and EMTREE databases, adapted for each database. There will be no restrictions on publication date or language. Descriptors will be combined using the Boolean operators ‘AND/OR’. It is worth noting that the descriptors and their synonyms were organized into sets and subdivided into lines, as shown in Table 1.

**Table 1:**
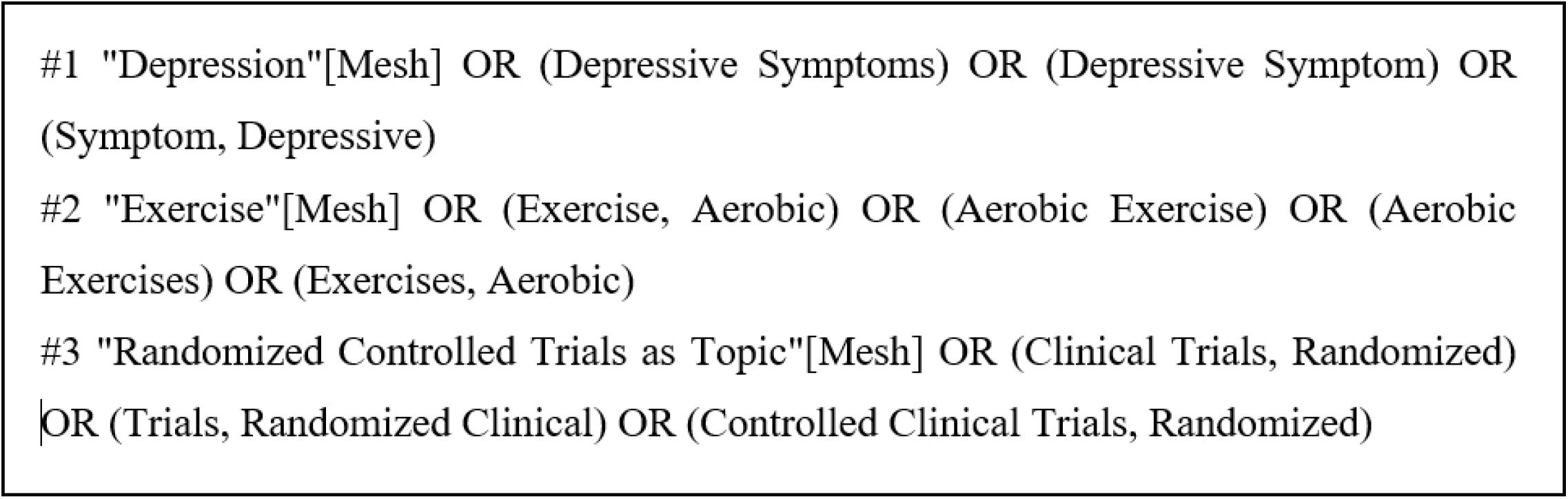
High-sensitivity search strategy.

This systematic review will follow the eligibility criteria based on PICO.

#### Participants

studies with adults (>18 years) of both sexes, without chronic degenerative diseases or diagnoses of more severe psychiatric disorders.

#### Intervention

aerobic exercises such as dancing, steps, walking, and running. Comparator: other interventions or control group.

#### Outcomes

depressive symptoms as the primary outcome. Training volume and frequency will be considered as secondary outcomes. Studies which used validated depression scales will be included, such as the Beck Depression Inventory (BDI), Hamilton Depression Rating Scale (HAM-D), Patient Health Questionnaire (PHQ-9), Center for Epidemiologic Studies Depression Scale (CES-D), Montgomery-Asberg Depression Rating Scale (MADRS), Zung Self-Rating Depression Scale (Zung SDS), Geriatric Depression Scale (GDS), and Hospital Anxiety and Depression Scale (HADS), among others.

#### Study type

randomized clinical trials. Only original studies will be eligible for inclusion in this study; ongoing clinical trials with preliminary results will not be included. Studies involving populations with non-communicable chronic diseases (i.e. patients with hypertension, diabetes), case reports, preprints, narrative and systematic reviews, and observational studies will be excluded.

After conducting a high-sensitivity search in the databases, articles will be selected using Rayyan software, a tool specialized in screening studies for systematic reviews and meta-analyses [5]. Two reviewers (L.N.S; M.I.K) will initially be registered on the platform, where they will receive the articles identified by the search. The technical assessment will be divided into two phases: both reviewers will examine the articles in the first phase to determine their inclusion in the review. A third senior and expert reviewer (I.K.S) will review any articles in case of disagreement, and make the final decision on its inclusion.

After applying the eligibility criteria, the selection process will be conducted in two stages. The titles and abstracts of the articles will be read in the software in the first stage, which must be accepted by at least one of the reviewers for it to proceed to the subsequent phases. Next, the articles will be read in full in the second stage, and acceptance by both reviewers will be required for them to be included in the study.

### Data extraction

The following data for each study included in the systematic review will be extracted in detail: bibliographic information (author names, study title, year of publication); sample characteristics (size, inclusion and exclusion criteria, as well as other relevant participant characteristics, such as physical condition); and methodological details (study type, description of the intervention and control groups, with an emphasis on the duration, intensity, and frequency of the exercises, as well as the scales used to measure the outcomes).

Primary and secondary outcomes will be analyzed focusing on the following aspects: changes in depressive symptoms as primary outcomes; training volume and activity frequency as secondary outcomes. The validated instruments used to assess these outcomes will also be recorded.

Quantitative results will include effect size, measures of variability such as confidence intervals, standard deviation, and p-values, as well as other data relevant for interpreting the results. When applicable, qualitative information such as emerging categories and themes will also be extracted. Finally, the authors’ main conclusions will be considered and recorded.

After collecting the described data, a synthesis table will be prepared to group the extracted information, including year of publication, authors, methods, interventions, and main results of each study. This table will facilitate a comparative analysis of the included studies and provide a clearer view of the reported outcomes.

Data for studies reporting multiple outcomes or using different measures for the same outcome will be extracted separately and treated as independent entries, thereby avoiding duplication of results. When more than one scale is used to assess depressive symptoms, the most widely used scale (i.e. the Hamilton Scale) will be prioritized, with a clear justification for the choice.

### Risk of bias assessment

The assessment of the methodological quality of the studies selected during screening will be conducted using the Risk of Bias 2.0 (RoB 2.0) tool [6]. The methodological quality will be analyzed by two reviewers (L.N.S; M.I.K). This method evaluates the quality of randomized clinical trials based on various criteria: randomization, intervention assignment, adherence to the intervention, missing outcome data, outcome measurement, selection of reported outcomes, and overall risk.

### Certainty of the evidence

The Grading of Recommendations Assessment, Development, and Evaluation (GRADE) is a widely used system for the methodological assessment of scientific studies, aimed at determining the level of evidence and strength of recommendations. This system is based on rigorous criteria to ensure the quality and reliability of the presented evidence. Fundamental requirements for applying GRADE include assessing the certainty of evidence, which determines the degree of confidence in the results, and analyzing specific domains such as risk of bias, inconsistency, imprecision, and potential publication bias. Additionally, it is important that the results are clear and appropriate. GRADE categorizes both the level of evidence and the strength of recommendations into four categories: high, moderate, low, and very low [7].

### Data synthesis

The meta-analysis will be conducted using a random-effects model, considering the expected variability among the included studies due to the heterogeneity of aerobic exercise modalities, intensities, frequencies, and durations assessed. This model is most appropriate when assuming that true effects vary across studies, allowing for a more robust and generalizable synthesis of results.

The random-effects model will be applied to all analyses, reflecting the assumption that true effects vary between studies due to differences in interventions and populations. The variance estimate for binary outcomes (i.e. presence/absence of depressive symptoms) across studies will be based on the Mantel-Haenszel method. The inverse variance method will be used for continuous outcomes (such as depression scores).

Heterogeneity among studies will be assessed using the I² index, which quantifies inconsistency among study results, and Cochran’s Q test, which evaluates the statistical significance of heterogeneity. If heterogeneity exceeds 80%, subgroup analyses will be conducted to investigate potential sources of variation, such as exercise intensity, study duration, or participant characteristics. If the cause of heterogeneity cannot be identified, the meta-analysis may be replaced by a narrative synthesis of the results. Review Manager software [8] will be used to conduct the statistical analyses. One review author will be responsible for entering data into the software, while a second author will check data accuracy.

### Subgroup analyses

Subgroup analyses will be conducted to evaluate the effectiveness of aerobic exercise in different contexts:

- Exercise intensity: classification based on perceived exertion rate or heart rate (e.g., light, moderate, vigorous).
- Intervention duration: comparison between short and long-duration interventions.
- Type of exercise: differentiation among aerobic exercise modalities (walking, running, cycling, etc.).
- Participant characteristics: analyses based on age groups (i.e. adults >18 years), gender, and mental health status.

Subgroup analysis is essential for exploring the effectiveness of aerobic exercise in different contexts and populations. Different exercise intensities (light, moderate, vigorous) may have variable effects on depression, and this differentiation will help identify the most effective modalities. Intervention duration can significantly influence outcomes, making it important to compare short- and long-duration interventions. Participant characteristics, such as age and gender, may also moderate the effects, justifying separate analysis for these groups.

### Sensitivity analysis

A sensitivity analysis will be conducted to assess the robustness of the results in relation to the risk of bias in the included studies. Studies with a high risk of bias will be identified, considering the worst judgment in any of the assessed domains, excluding blinding domains due to the nature of physical exercise interventions. Excluding these studies will allow us to evaluate whether they significantly influence the overall results of the meta-analysis.

### Strategy for handling missing data

Data imputation methods or intention-to-treat analysis will be applied to handle missing data, ensuring that the meta-analysis results are complete and representative.

### Publication bias

Publication bias will be examined through funnel plots and Egger’s test. If significant bias is detected, correction techniques, such as the trim-and-fill method, will be considered to adjust the results.

### Presentation of results

The results will be presented in forest plot graphs, which will illustrate the combined effects of the interventions and the heterogeneity among the studies. Additionally, a narrative synthesis will discuss the clinical implications of the findings, the limitations of the included studies, and the research gaps that need to be addressed in future investigations.

## Discussion

The literature widely documents the effectiveness of aerobic exercise for clinically improving adults with depression, although the topic still requires further exploration due to the complexity of the variables involved. Aerobic exercise has shown results in reducing depressive symptoms across different populations, being comparable to various therapeutic approaches, including medication, psychotherapy, and other exercise-based therapies. Numerous studies seek to evaluate this relationship, as exemplified in Olson et al. [9], which reveals that moderate-intensity aerobic exercise not only reduces depressive symptoms, but also improves cognitive control, suggesting benefits beyond emotional relief. Similarly, a study focused on geriatric interventions demonstrated that short-duration aerobic exercise yielded promising results in hospitalized patients, indicating that even brief interventions can be effective [10].

A detailed analysis of exercise intensity in studies indicates that while more intense activities may offer additional benefits, effectiveness in reducing depressive symptoms is not limited to high-intensity levels. Recent research demonstrates that both light and intense exercises promote relief of depressive symptoms, with no significant differences between groups of varying intensities [11,12]. This suggests that exercise intensity can be adapted according to individual needs for initial management of depression.

Given the gaps in the literature that have yet to establish an ideal dose-response for physical exercise, a clinical trial evaluated the impact of different intensities (light, moderate, and vigorous) on depressive symptoms. The results showed a significant reduction in symptoms across all groups, with no statistically relevant differences between intensities. This data underscores the need for comprehensive systematic reviews, like the present one, to identify the most effective combination of exercise intensity and volume, considering the specificities of different clinical profiles [11].

In summary, the benefits of exercise go beyond improvements in mental health parameters; research reveals connections with cognitive and physiological aspects. A study comparing aerobic exercise with stretching, for instance, showed improvements in visuospatial memory and cardiorespiratory capacity in individuals who engaged in aerobic exercise [13]. These results paradoxically emphasize what existing literature suggests: physical activity can promote neurobiological changes, such as increased neuroplasticity and enhanced cognitive function.

Given the challenges of adherence to exercise programs among individuals with depressive characteristics, the ‘DEMO-II’ study, for example, encountered difficulties in maintaining high participation levels, which may have limited the results [13]. To overcome this, strategies which consider patients’ intrinsic motivation and preferences are essential. Low-intensity exercises, which are less strenuous, may be appealing to increase adherence and ensure continuous treatment.

The study results indicate that exercise should be considered an effective complement to depression treatment. Research evaluating the efficacy of exercise as an addition to standard treatment revealed that it can enhance the effects of traditional interventions, promoting improvements in quality of life and sustained reductions in depressive symptoms. Although exercise is not yet considered an essential component on its own, the findings suggest it has great potential to be integrated as a significant part of therapeutic strategies, especially for patients who do not fully respond to conventional treatments [14].

The reviewed studies present some methodological limitations, such as small sample sizes, a lack of clarity on the specific mechanisms through which exercise contributes to improvements in depression, and variation in the exercise intensities considered most effective. However, this review aims to precisely highlight these conclusions and shed light on these issues.

## Conclusion

The systematic review of RCTs aims to present findings on reducing depression levels in adults and to identify the most effective dose-response for this reduction, specifying which intensities, frequencies, and durations of aerobic exercise produce the most favorable outcomes. Additionally, the review is expected to offer in-depth understanding of the biological and psychological mechanisms through which aerobic exercise influences depression levels.

## Data Availability

N/A - Not applicable

## Author Contributions

Conceptualization: Larissa Nayara de Souza; Silvana Medeiros de Araújo; Eva da Silva Paiva; Alícia Eliege da Silva; Joel Florêncio da Costa Neto ; Juvêncio César Lima Assis ;Isis Kelly dos Santos ; Themis Cristina Mesquita Soares; Edson Fonseca Pinto ; Roque Ribeiro da Silva Júnior; Maria Irany Knackfuss.

Data curation: Larissa Nayara de Souza; Silvana Medeiros de Araújo; Eva da Silva Paiva; Alícia Eliege da Silva; Joel Florêncio da Costa Neto; Juvêncio César Lima Assi and Maria Irany Knackfuss.

Formal analysis: Larissa Nayara de Souza; Isis Kelly dos Santos; Themis Cristina Mesquita Soares; Edson Fonseca Pinto; Roque Ribeiro da Silva Júnior; Maria Irany Knackfuss.

Funding acquisition: Maria Irany Knackfuss.

Investigation: Larissa Nayara de Souza; Silvana Medeiros de Araújo; Eva da Silva Paiva; Alícia Eliege da Silva; Joel Florêncio da Costa Neto; Juvêncio César Lima Assi and Maria Irany Knackfuss.

Methodology: Larissa Nayara de Souza; Isis Kelly dos Santos; Themis Cristina Mesquita Soares; Edson Fonseca Pinto; Roque Ribeiro da Silva Júnior; Maria Irany Knackfuss.

Funding acquisition: Maria Irany Knackfuss.

Project administration: Larissa Nayara de Souza; Isis Kelly dos Santos and Maria Irany Knackfuss.

Supervision: Isis Kelly dos Santos and Maria Irany Knackfuss.

Validation: Larissa Nayara de Souza; Isis Kelly dos Santos; Themis Cristina Mesquita Soares; Edson Fonseca Pinto; Roque Ribeiro da Silva Júnior; Maria Irany Knackfuss.

Visualization: Larissa Nayara de Souza.

Writing – original draft: Larissa Nayara de Souza; Silvana Medeiros de Araújo; Eva da Silva Paiva; Alícia Eliege da Silva; Joel Florêncio da Costa Neto; Juvêncio César Lima Assi and Maria Irany Knackfuss.

Writing – review & editing: Larissa Nayara de Souza; Isis Kelly dos Santos; Themis Cristina Mesquita Soares; Edson Fonseca Pinto; Roque Ribeiro da Silva Júnior; Maria Irany Knackfuss.

---

**Fig 1:**
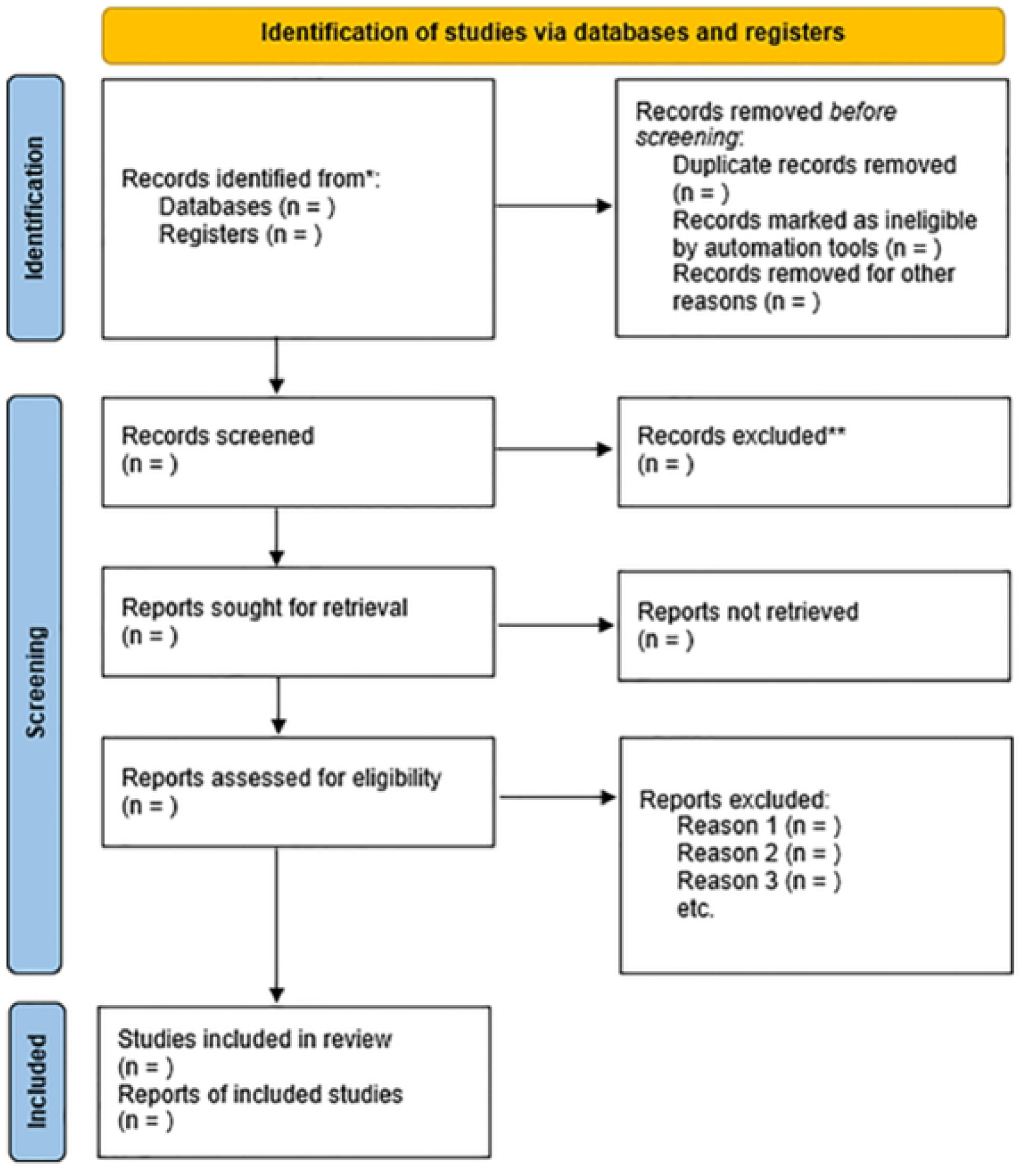
PRISMA flow diagram for systematic review and meta-analysis

